# Technical and scale efficiency in health service production in Kenya: subnational analysis of 47 county governments in 2022

**DOI:** 10.1101/2025.08.02.25332872

**Authors:** Tom Achoki, Naomi Rotich, Dellany K. Bett, Josephat Tonui, Tabitha Oanda, Abaleng Lesego, Richard Wamai, Yohannes Kinfu, Uzma Alam, Walter Obiero, Lawrence P.O. Were, Matthew Schneider

## Abstract

**Introduction:** Kenya adopted a decentralized health system as part of the efforts to improve access, eliminate inequalities and make progress towards universal health coverage (UHC). With significant investments, county governments are charged with the responsibility of delivering healthcare to their population. However, questions remain about the efficiency of resource utilization to meet the health goals.

**Methods:** We assembled a dataset comprising health sector inputs, outputs, and contextual factors to measure the efficiency of Kenya’s health system across 47 counties in 2022. We estimated overall technical efficiency (OTE), pure technical efficiency (PTE) and scale efficiency (SE) employing Simar and Wilson’s single-step data envelopment analysis (DEA) approach. We assessed the impact of contextual factors on efficiency levels using a truncated regression model.

**Results:** Considering child survival as a health system output, the national average for OTE was 0.74 (95% CI:0.70-0.78), while PTE was 0.85 (95%CI:0.82-0.88) and SE was 0.87 (95%CI:0.85-0.89). Meanwhile, for childhood immunization coverage, average OTE was 0.83 (95%CI:0.81-0.87), while PTE was consistently high across the counties. For maternal survival, OTE was low at 0.51 (95%CI:0.48-0.55), and PTE was 0.61 (95%CI:0.57-0.69), with SE averaging 0.82 (95%CI:0.79-0.84). Taking healthy adjusted life expectancy (HALE) as the health system output, average OTE was 0.83 (95%CI:0.81-0.85). There was a high correlation between OTE scores that considered child survival, childhood immunization coverage and HALE as health system outputs. Efficiency scores showed a heterogenous picture across the country both at the provincial and county levels.

**Conclusion:** In 2022 the Kenyan health system was inefficient. Decision makers in Kenya have an opportunity to improve health outcomes without the injection of additional resources particularly through better managerial practices as pointed by low attainment in PTE. Additionally, reorganizing the scale of health programs to operate at the optimal level would yield improved efficiency.

## 1. Background

Kenya’s successive governments have considered achieving the highest possible health as an ultimate indicator of—and a foundation for—national development.^1^ ^2^ Several government programs, policies and development plans, including Vision 2030, prioritized improving and expanding health service delivery.^3^ Since 2010, the government has adopted a two-tier health governance structure that delegated service delivery to the 47 autonomous popularly elected subnational governments, known as counties.^4^ The counties were determined based on historical-political antecedents following the first post-colonial Constitution passed in 2010 rather than on any geographical or economic basis.^5^ The new governance structure tasked the central government with setting norms, standards, and guidelines for the entire health system.^6^ This includes policy formulation, managing referral hospitals, coordinating national health programs, mobilizing domestic and external resources, undertaking supervision, capacity building, and technical assistance to counties when required. The central government also allocates health resources to counties according to population size and pre-existing inequalities within counties.^7^

On the other hand, counties are responsible for planning, organizing, and providing health services and interventions within their jurisdictions. They also play a crucial role in implementing Kenya’s landmark goal of achieving UHC by 2030.^8^ ^9^ This allowed the local governments to invest heavily in constructing new health facilities and using resources to recruit new medical staff, procure locally required medications, and purchase new medical equipment and ambulances.^10^ They have also adopted health initiatives tailored to their populations, including mobilizing local efforts and resources to address environmental and socioeconomic factors that have a bearing on population health.^11^

Implicit in these new investments is the expectation by decision-makers that health systems are already efficient, meaning that there is limited space for improving health goals by enhancing inputs’ efficiency or reallocating them differently for better results. However, studies in Africa and elsewhere quantified that technical and allocative inefficiencies are widespread and, in most cases, closely associated with geographical disparities in service coverage and health outcomes.^12^ For example, a district level study in Zambia demonstrated that the country’s health system was functioning less than 62 % of its optimal capacity, pointing to the potential for expanding services without injecting additional resources into the system.^13^ Kinfu and Sawhney’s^14^ and Chai et al^15^ showed geographical disparities in technical and allocative inefficiency in maternal care and NCD service delivery in India and China. Several studies have revealed low technical efficiency in various HIV interventions in Nigeria, South Africa and Rwanda.^16^

Overall, in Kenya, the health system reform and the health sector investment return over the past few years have been considerable.^17^ Nationally, life expectancy at birth increased by 5·4 years (95% UI 3·7– 7·2) between 1990 and 2016: from 61·4 years (95% UI 60·8–62·0) in 1990 to 66·8 years (66·1–67·6) in 2016. Reductions in maternal mortality were equally notable, although the existence of substantial heterogeneity revealed relevant performance gaps that needed addressing. The same applies to under-5 mortality, with some counties showing annual declines of greater than 5%, whereas, in others, the level remained stable over time. Besides, even in the counties which achieved better outcomes, the recorded gains were significantly lower than those attained in other countries with comparable development levels.^18^ These raise several important health system performance questions that we endeavor to address.

There have been a considerable number of studies on Kenya’s health system performance, but they have notable limitations.^19^ First, the analyses were focused on selected programs or health facilities rather than on the entire health system.^20^ Second, efficiency was measured in these studies using healthcare activities or sheer production of services.^21^ However, activity (or service volume) based efficiency analyses assume homogeneity in service quality and disregard adverse effects on patients or care providers.^22^ Our study mitigates these shortcomings in three fundamental ways. First, we used health outcomes rather than activities as a measure of health systems output. Second, we employed a county-level health system analysis rather than a narrow health facility-level efficiency analysis. Thirdly, we measured health system performance at the county level, primarily because counties are responsible for planning, organizing, and providing health services and interventions in Kenya.^23^ To our knowledge, this is the first study in Kenya to take such a holistic view in assessing the efficiency of the subnational governments since they were created in 2013.

This study becomes fundamentally important particularly at a time when shifts in global funding and reduced donor support expose the vulnerability of national health systems. Top in the minds of decision makers is whether Kenya can achieve better outcomes with the resources already invested in the health system? Second, which counties are technically inefficient and what factors account for the differences? Finally, what implications can we draw from our findings towards building a resilient health system that attains UHC by 2030?

## 2. Methodology

### 2.1. Efficiency analysis

Health efficiency analyses distinguish between technical, allocative and scale efficiency and between input and output orientations. ^24^ Because input prices needed to estimate allocative efficiency were unavailable in our case, we only considered technical and scale efficiencies. We employed an output rather than an input-oriented efficiency model because health systems face a fixed set of core health inputs in the short run, and decision makers can only organize the utilization of the available resources to achieve certain outcomes. Therefore, an output-oriented model was justified because health systems focus on achieving the highest possible outcome, given their resources, rather than setting a target outcome level and removing resources once they attain their targets.

We measured technical efficiency using DEA, a non-parametric method based on linear programming that can handle a multidimensional mix of input and output variables.^25^ The Charnes, Cooper and Rhodes (CCR)^26^ and the Banker, Charnes and Cooper (BCC)^27^ are the most extensively used models within the DEA framework. The CCR model assumes constant returns to scale, while the BCC assumes variable returns to scale. Technical efficiency scores obtained using the CCR model represent overall technical efficiency (OTE), while those obtained using the BCC model represent pure technical efficiency (PTE). Dividing OTE by PTE provides another important measure of scale efficiency (SE).^28^

Briefly, OTE measures how well a decision-making unit (DMU) generally transforms inputs into outputs. In this respect, a DMU is technically efficient if it operates on the efficient frontier, using the minimum possible inputs to produce a given level of output. Meanwhile, PTE measures the efficiency of a DMU in converting inputs into outputs regardless of its size or scale of operations. This means, it isolates managerial performance from scale effects. Therefore, a DMU with high PTE uses its resources efficiently, even if it’s not operating at the optimal scale. SE, on the other hand measures whether a DMU is operating at an optimal scale. ^28^ ^29^

We generated all three measures at the county level accounting for the different health system outputs under consideration given that the Kenyan health system is largely organized vertically following different health programming areas.^30^ To give further context to the county level estimates, we also grouped the counties into the provinces (regions) in which they were previously clustered. Since Nairobi County was classified as a province, we group it into the Central province given its proximity to the region.

Algebraically, DEA is achieved by solving for each DMU, which in our case is the county, the following linear programming problem.

Maximize θ

subject to:

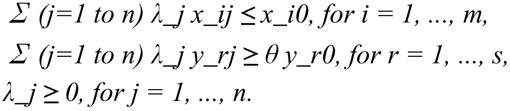

where:

– θ is the efficiency score (output expansion factor),
– x_ij and y_rj are the inputs and outputs for DMU j,
– x_i0 and y_r0 are the inputs and outputs for the DMU being evaluated,
– λ_j are the weights assigned to each DMU.

We adopted Simar and Wilson’s (S-W) approach to estimate the efficiency indices, which provides bias-corrected efficiency scores and their determinants in a single step, using a bootstrapping technique. ^31^ The advantage of the S-W approach is that the estimation process does not impose a functional form on the efficiency term; hence it is non-parametric.^32^ Finally, we applied a truncated regression technique embedded within the S-W framework to control for contextual/environmental factors influencing efficiency scores. We used bootstrap procedures to generate bias-corrected confidence intervals for our estimates. ^33^ ^34^

### 2.2. Data and variable description

Three sets of data are required to perform efficiency analysis: data on health sector inputs, health outputs, and contextual or environmental factors. For our case, we used population adjusted health workforce and the development health budget as health systems inputs. Health system outputs included child immunization coverage, HALE, probability of survival for under-five year olds and maternal survival. Under-five year old and maternal survival were calculated as the reciprocal of under-five mortality rate (U5MR) and maternal mortality rate (MMR) for each county. In Kenya, health inputs are generally under the direct control of counties.^35^ However, each county faces a heterogeneous environment beyond their control that may positively or adversely affect their ability to combine inputs efficiently. For instance, well-developed counties are likely to have a well-educated population with better access to social and electronic media.^36^ Such populations are likely to be better informed on health matters and likely to have the resources to purchase health-enhancing commodities, which will enable the county to achieve higher efficiency for health outcomes without any additional health input of its own. Therefore, for environmental factors, we considered women education, access to water and access to electricity as determinants of health performance.

County level data were assembled from multiple sources available for the year 2022. Health workforce and the development health budget data were obtained from the surveys conducted by the Kenya National Bureau of Statistics (KNBS).^37^ ^38^ Similarly, data on childhood immunization coverage, women education, access to water and electricity were obtained from the Kenya Demographic and Health Survey of 2022, that was conducted by the KNBS. HALEs, under five mortality rate and maternal mortality rate were obtained from the Global Burden of Disease Study subnational estimates for Kenya, produced by the Institute for Health Metrics and Evaluation (IHME) at the University of Washington.^39^ Each health system output was considered separately in the efficiency analysis, using the same inputs and environmental variables to gain insight into specific aspects of the health system production. All variables were log-transformed to mitigate skewness and stabilize variance, which improves the robustness of DEA and regression estimates.

### 2.3. Data analysis and management procedures

We used Stata version 16.1 for data management procedures, technical analyses and visualizations (StataCorp. 2019. Stata Statistical Software: Release 16.1 College Station, TX: StataCorp LP.^40^ For S-W analysis we used R-software applying the Robust Data Envelopment Analysis (r-DEA) package and ran 10000 bootstrap replications to generate bias-corrected efficiency measures. The number of replications in our analysis is well above the level recommended by S-W.^41^

#### Ethical approval

Permission to conduct the study was granted by the Ministry of Health in Kenya and the respective national institutions that provided the data used on the analysis. Since our study used only deidentified publicly available secondary data at the county level, it was exempt from the full institutional board review.

## Results

### Descriptive statistics

Table 1 shows the descriptive statistics for the inputs, outputs and environmental variables that were used in the study. Several variables showed wide variation across the 47 counties. For instance, the development health budget to counties averaged 53.92 million Kesh/100,000 population, with a range of between 10.40 million Kesh/100,000 population and 196.12 million Kesh/100,000 population, while health workforce density averaged 16.40 health workers/ 10,000 population with a range of 6.75 health workers/10,000 population and 54.74 health workers/10,0000 population. Similarly, U5MR averaged 42.53 deaths/ 1000 live births, but ranging between 15.00 deaths/1000 live births and 73.00 deaths/1000 live births. MMR also had a wide range of between 64.66 deaths /100,000 live births and 855.95 deaths /100,000 live births. Environmental variables also showed similar heterogeneity across counties, with household access to electricity, ranging between 16.30% and 94.50%, while household access to a clean water source ranged between 22.80% and 89.70%.

**Table 1:**
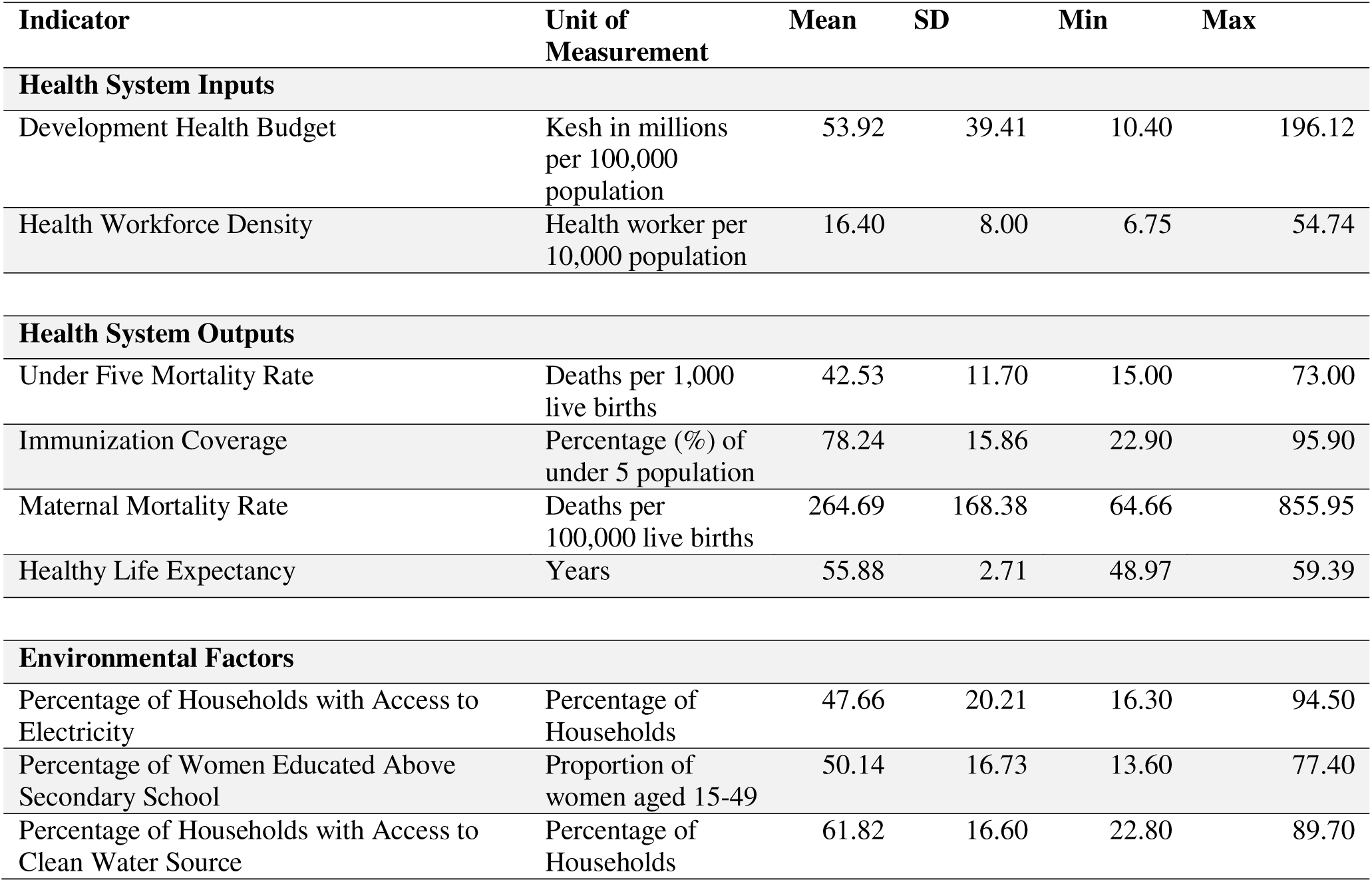
Summary of Variables.

### Overall efficiency, pure technical efficiency and scale efficiency

Figure 1 shows the OTE, PTE and SE across the 47 counties in Kenya, when considering child survival as the health system output variable. Only 14 counties out of the 47 (approximately 30%), had OTE equal or above 0.80; with most of the low performance concentrated in counties in Central, Coast, Rift Valley and Nyanza provinces. Considering PTE, 35 counties (approximately 75%) scored 0.80 and above, and low performance was mainly clustered in northern parts of the Rift Valley, Nyanza and Western provinces. Similarly, SE was high across majority of counties, with low performance in parts of Eastern and Rift Valley provinces.

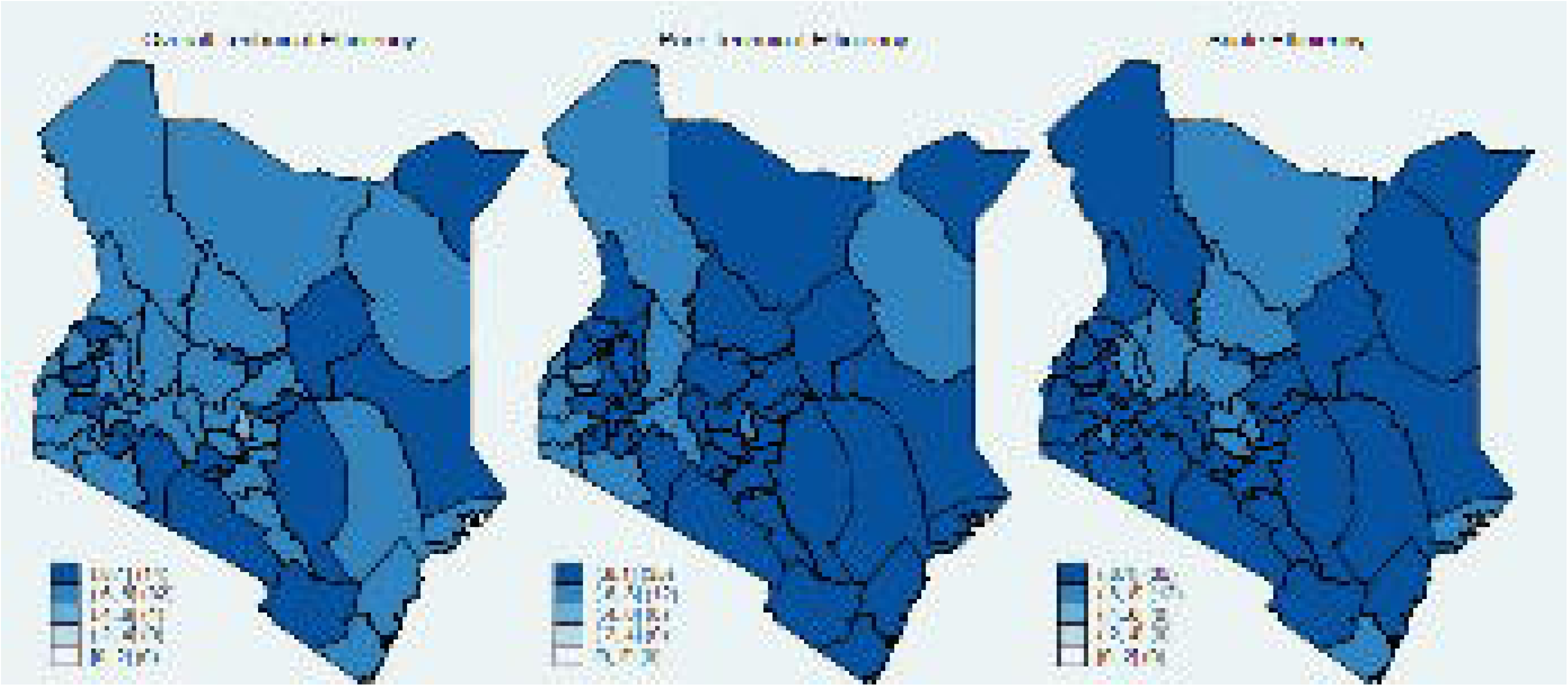

Appendix 1 which focuses on child survival, ranks the counties from the best performing in terms of OTE, which is Bomet, at 0.94 (95CI: 0.89 – 1.00) to the worst performing, Kirinyaga at 0.59 (95% CI: 0.56-0.62). The national average for OTE was 0.74 (95% CI:0.70-0.78). Meanwhile, in terms of PTE, Tharaka Nithi and Mandera were leading the pack at 0.96 (95%CI: 0.93-0.99) and 0.96(95%CI: 0.94-1.00) respectively. The national average for PTE was 0.85 (95%CI:0.82-0.88). On the other hand, the national average for SE was 0.87 (95%CI:0.85-0.89). Several counties including Bomet, Kitui, Kiambu and Kakamega performed highly averaging 1 in terms of SE, which is fully scale efficient.

Figure 2 shows the county level distribution of OTE, PTE and SE in terms of childhood immunization coverage. Considering OTE, only 32 counties (approximately 68%) had a score above or equal to 0.80, with low performance clustered in the Coast, Eastern, parts of Rift Valley and the Western provinces. All counties scored above 0.80 in terms of PTE, while SE, was low in parts of Central and Eastern provinces.

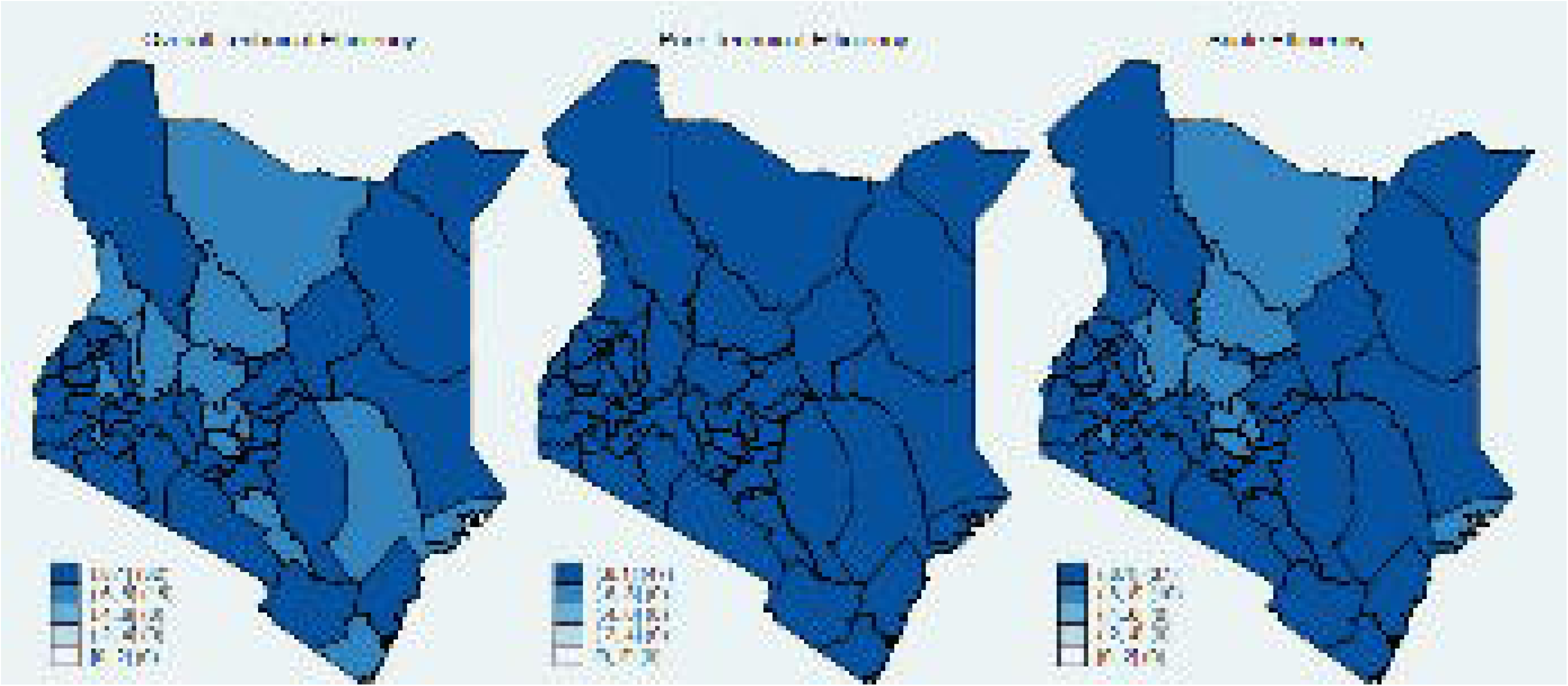

Appendix 2, which considers childhood immunization as a health system output, ranks the county level OTE from the best performers, which are Bungoma 0.97 (95% CI:0.95-1.00), Nairobi 0.97 (95%CI: 0.94-1.00) and Mombasa 0.97 (95%CI:0.94-1.00), to the worst performance of Elgeyo Marakwet 0.7 (95%CI:0.68-0.72) and Marsabit 0.69 (95%CI: 0.68-0.72). The national average for OTE was 0.83 (95%CI:0.81-0.87), while PTE was consistently high across the counties with a national average of 0.96 (95%CI: 0.95-0.97). SE showed variation across the counties, mirroring the observed distribution pattern of OTE, particularly in the Eastern province. The national average was 0.87 (95%CI:0.85-0.89)

The efficiency indicators regarding county level maternal survival are shown on Figure 3. All indicators show considerable heterogeneity across the 47 counties. Only 3 counties had an OTE score above or equal to 0.80, while only 10 had the same for PTE. In terms of OTE and PTE, low performance was widespread particularly in parts of the Central and Eastern provinces, however there was no clear regional trend.

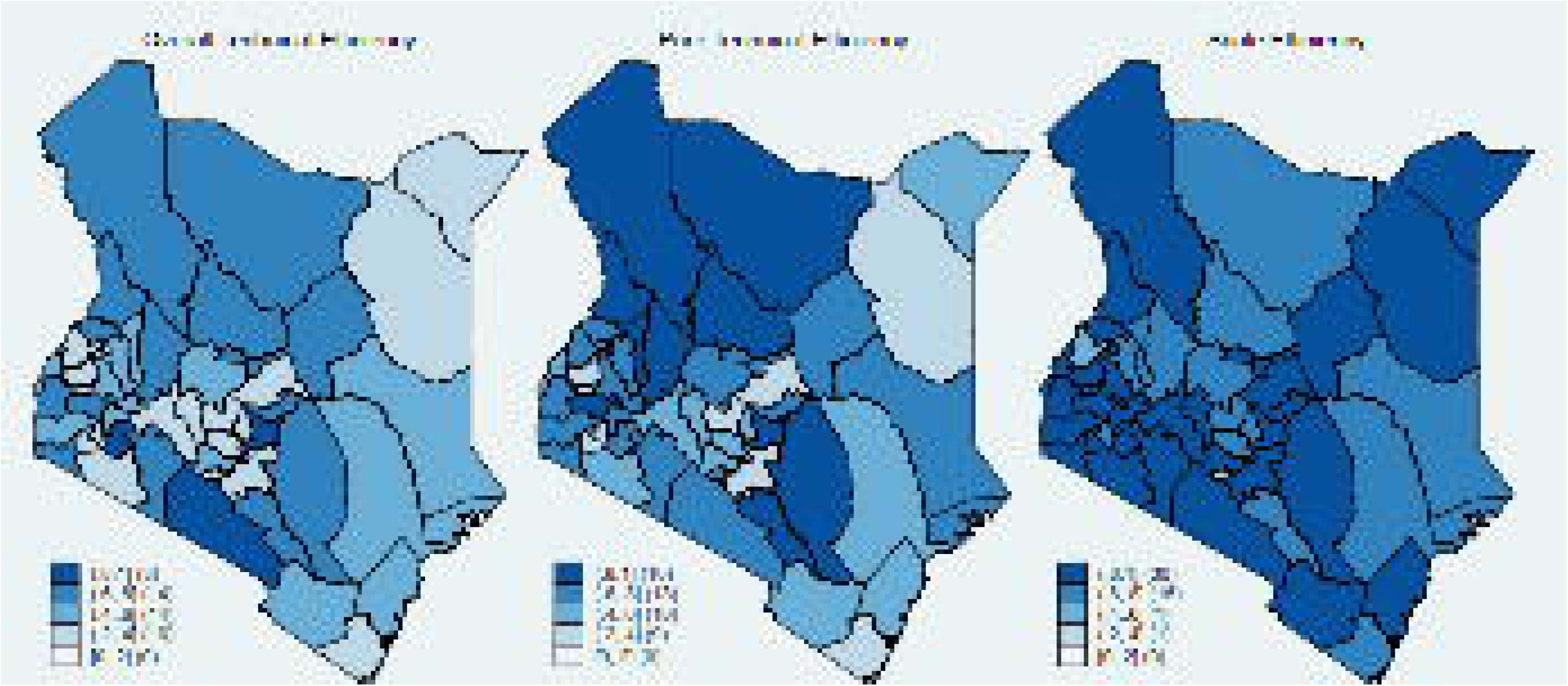

Appendix 3, which considered maternal survival as a health system output ranks the counties from the best performers in terms of OTE, Bomet at 0.94 (95%CI: 0.89-0.99) followed by Kajiado at 0.89 (95%CI:0.83-0.98), to the worst performing; Kisii at 0.22 (95%CI:0.21-0.24) and Nyeri at 0.21 (95%CI:0.20-0.22). The national average OTE was low at 0.51 (95%CI:0.48-0.55). PTE also showed wide variation across the country with the top performing counties being, Marsabit 0.98 (95%CI: 0.95-1.00), Samburu 0.97 (95%CI:0.94-1.00) and West Pokot 0.96 (95%CI:0.93-1.00). The national average PTE was 0.61 (95%CI:0.57-0.69). The national average SE was generally high at 0.82 (95%CI:0.79-0.84), with a heterogenous picture across the country. Generally, the counties that had high OTE also performed well in terms of SE, with a few exceptions, such as Kakamega, Kiambu, Isiolo and Tharaka Nithi. These four counties were fully scale efficient but performed poorly in terms of OTE.

Figure 4 shows the efficiency indices considering HALE as the main health system output. Only 28 counties (approximately 60%) had OTE above or equal to 0.80. Low performance was concentrated in the Central, Coast, Eastern, parts of Rift Valley and Nyanza provinces, somewhat overlapping with trends observed in child survival. A similar pattern was observed for SE, with only 31 counties (approximately 66%) having a score above or equal to 0.80. Meanwhile, PTE was uniformly high across the country.

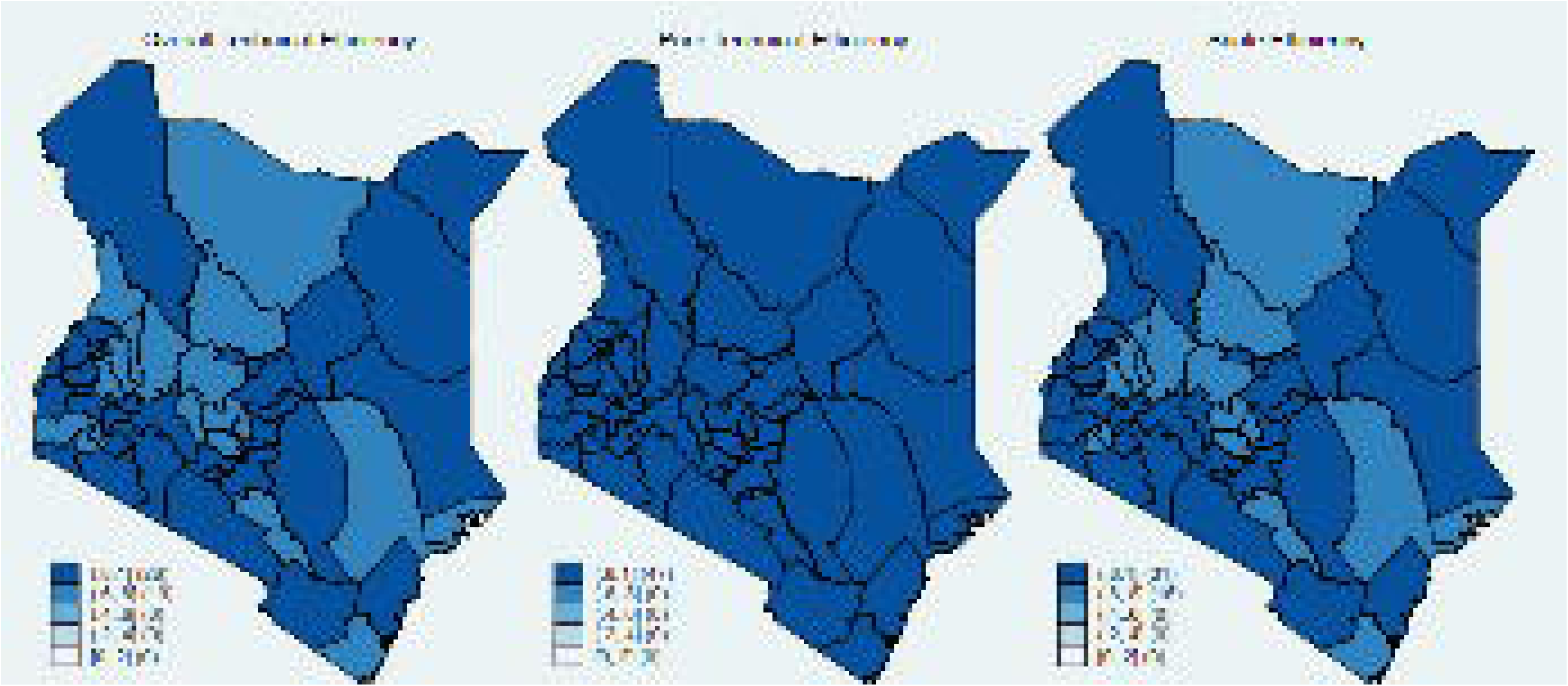

Appendix 4, which considers HALE as the health system output shows the ranking of counties in terms of OTE from the best performers, Kitui 0.97 ( 95%CI:0.96-1.00) and Bungoma 0.97 (95%CI: 0.95-1.00) to the worst performers, Nyamira 0.71 (95%CI:0.69-0.73), Lamu 0.71 (95%CI:0.69-0.73) and Marsabit 0.70 (95%CI:0.69-0.72). The national average OTE was 0.83 (95%CI:0.81-0.85). SE followed a similar pattern as OTE with considerable heterogeneity across the country. The average SE was 0.84 (95%CI:0.82-0.86), while PTE was uniformly high across the counties averaging 0.99 (95%CI:0.98-0.99).

Appendix 5 shows the heterogeneity of the country’s health system performance at the provincial or regional level in terms of OTE. In terms of OTE for child survival, it shows Eastern province leading and Nyanza province trailing. The Western province leads when considering childhood immunization as a health system output while Rift Valley leads in maternal survival. Meanwhile, the Western province leads when considering HALE as the health output while Rift Valley trails.

Figure 5 shows how the OTE scores resulting from the different health system output variables are correlated. There is a strong correlation between OTE scores from immunization and child survival, r=0.74. Similarly, there is a strong correlation between the OTE scores resulting from immunization and HALEs, r=0.96, as well as OTE scores from child survival and HALEs, r=0.81.

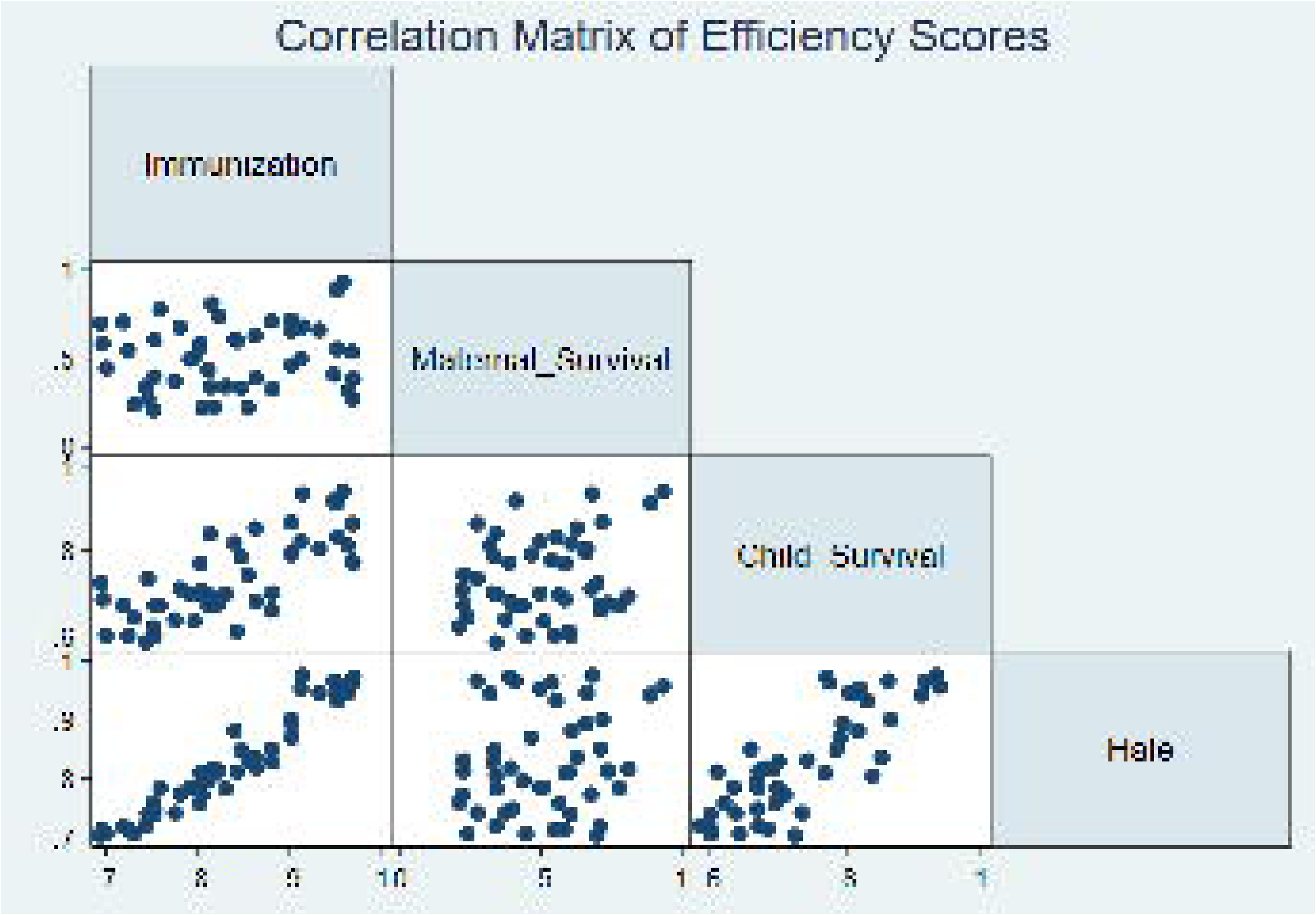

### Environmental factors

Table 2 shows the truncated regression results for factors beyond the healthcare system. In our model, the dependent variable is the reciprocal of the efficiency score; meaning if efficiency is E, then the reciprocal is (1/E). Therefore, for each unit increase in the log of access to clean water, the reciprocal efficiency score related to child survival decreases by 0.08 units. Similarly, for each unit increase in the log of access to electricity, the reciprocal efficiency score related to child survival decreases by 0.05 units. Since the negative coefficient means the reciprocal efficiency score decreases, the actual efficiency score increases by the same magnitude. This signifies a positive relationship, but both are not statistically significant since the confidence intervals contain 0. Women education increase also had a positive relation with the efficiency score related to maternal survival with a unit increased in its log leading to a 0.63 decrease in the reciprocal of the efficiency score, however the relationship was not statistically significant.

|  | Child survival |  |  | Immunization |  |  | Maternal survival |  |  | Hale |  |  |
| --- | --- | --- | --- | --- | --- | --- | --- | --- | --- | --- | --- | --- |
|  | coeff | CI.2.5. | CI.97.5. | coeff | CI.2.5. | CI.97.5. | coeff | CI.2.5. | CI.97.5. | coeff | CI.2.5. | CI.97.5. |
| Intercept | 1.72 | 0.90 | 2.60 | 1.70 | 1.04 | 2.38 | -7.21 | -17.79 | 12.76 | 1.54 | 0.84 | 2.28 |
| Women education | 0.05 | -0.15 | 0.23 | -0.07 | -0.22 | 0.06 | -0.63 | -3.30 | 2.36 | -0.07 | -0.23 | 0.09 |
| Access to water | -0.08 | -0.31 | 0.13 | -0.05 | -0.23 | 0.11 | 0.94 | -3.21 | 3.49 | -0.02 | -0.20 | 0.16 |
| Access to electricity | -0.05 | -0.20 | 0.11 | -0.01 | -0.13 | 0.12 | 1.81 | -1.17 | 3.81 | 0.00 | -0.14 | 0.13 |

## Discussion

Efficient use of available resources within the health system is central to making progress towards universal health coverage (UHC).^42^ This is particularly true in the current global health financing climate, with reduced development assistance for health. Decision makers in many resource-constrained countries like Kenya that have traditionally benefitted from the largesse of the donor community, particularly the US are now hard pressed to find sustainable ways to finance healthcare.^43^ ^44^ The US government retreat from funding global health initiatives has cited inefficiency and wastage of the US taxpayers’ money as one of the reasons for the abandoning its funding commitments.^45^ This comes at a time when health system decision-makers in many low- and middle-income countries are grappling with competing health priorities, including the recent infectious disease epidemics.^46^

Based on that context, we assessed Kenya’s health system’s technical and scale efficiency using recent cross-sectional data from 47 of its counties. Within Kenya’s decentralized health system, counties are responsible for planning and implementing health programs towards UHC, while policymaking and resource allocation are national level responsibilities.^8^ The analysis has further attempted to assess contextual factors explaining the county differences in health system efficiency. This analysis is particularly relevant given increasing evidence pointing to the fact that factors beyond the health system could significantly influence performance and overall population health outcomes. ^47^

Overall, the study reveals that Kenya’s health system was inefficient in 2022. There was an opportunity to further reduce U5MR from 42.53 deaths/1000 live births in 2022 by up to 24% without the injection of additional resources. We find that Kenya could reduce U5MR by 15% by improving administrative and organizational capacity at the county level health systems. There are considerable differences in managerial and organizational capacity between counties. For instance, in Migori county, U5MR could be improved by over 27% through enhancement of managerial capacity compared to the best performing county in the country-Mandera.

Our study reveals that the inappropriate size of county health operations (too large or too small) is also a source of inefficiency in the Kenyan healthcare system. On average, if all counties operated optimally, U5MR would reduce by 13.0%. Some of the counties are lagging in performance because of the inappropriate scale of their health operations. This type of inefficiency takes two forms: decreasing returns-to scale (DRS) and increasing returns-to-scale (IRS). DRS (also known as diseconomies of scale) implies that a county health system is too large to take full advantage of scale and has supra-optimum scale size. In contrast, a county experiencing IRS (also known as economies of scale) is too small for its scale of operations and, thus, operates at sub-optimum scale size.^48^ In our analysis most of the counties operated at IRS in the delivery of health services, indicating that additional inputs would have higher returns. Therefore, decision-makers at both national and county levels need to carefully examine the size and organization of the current county health systems and determine if they could be reorganized into optimal formations to maximize population health gains.

The country has the potential to improve childhood immunization coverage from its average of 78.24% by up to 17% without the injection of additional resources. Majority of the counties had good management practices with only a chance to improve coverage by a further 4% if all counties performed optimally in that respect. However, the greater opportunity for improvement lies in reorganization of the scale of childhood immunization operations, which could yield a further 13% improvement in immunization coverage without additional resources. Given that the counties are of different geographical sizes and population densities, decision makers might have to explore innovative ways to expand childhood immunization, in their respective counties.

MMR could be further reduced from its average of 264.69 deaths/ 100, 000 live births, by up to 49% without additional resources. Most of the improvements could be because of better management practices, with a potential to reduce MMR by approximately 39%, if all counties adopted optimal managerial practices. On the other hand, optimal reorganization of the scale of operations related to maternal survival would result in reductions of MMR of approximately 18%. Therefore, the efforts to improve health system performance should be tailored to specific county circumstances. For example, Marsabit and Samburu would only reduce MMR by 2% and 3% respectively through better managerial practices, while counties like Nyeri and Kisii have an opportunity to improve by 72% and 73% respectively by adopting optimal managerial practices. On the other hand, Marsabit and Samburu would make gains of approximately 28% and 27% respectively, by reorganizing the scale of their operations.

Considering, HALE as an overall health output indicator, Kenya has an opportunity for a further increase of 17% from its average of 55.88 years, without additional resources. Most of these gains would be through the reorganization of the scale of health delivery operations covering different areas, where approximately 16% gains would be realized. The fact that child survival (and by extension childhood immunization) which have been described above are significant contributors of HALEs means that the trends have a similar pattern. This is also illustrated by the strong correlations observed from the respective OTEs for the different health outputs considered in our analysis.

Further examination of subnational trends points to regional performance clustering whereby different regions are trading places when considering different health system outputs. This could be attributed to the influence of various factors, both internal and external, on the county. For instance, internal factors could include epidemiological trends of important diseases such as malaria, HIV/AIDS and non-communicable diseases and their risk factors.^49^ Similarly, demographic factors, socio-economic development levels and the geographical size of the county affect its population distribution and density concerning the available health system capacity.^50^ ^51^

Considering the importance of internal factors as determinants of health system efficiency, urban residents tend to have better access to health services, both in physical and financial terms, than their rural counterparts, resulting in better utilization of available services, which invariably translates to better health gains.^38^ ^52^ In contrast, due to access challenges in rural areas, there is often low utilization of essential health services. For example, child immunization rates for measles in the relatively urban and wealthier Nairobi and Mombasa counties are higher than the fairly rural Marsabit and Elgeyo Marakwet counties.^53^ Therefore, if significant progress is to be made, effective health programming cannot ignore important health determinants, such as socio-economic development, education, financial and physical barriers that ultimately determine health utilization and the accrued population health gains. Similarly, educated mothers with financial means are likely to be aware of and demand appropriate health services such as immunizations for their babies, increasing their odds of survival.^54^ This is particularly instructive for Kenya’s government’s recent efforts to improve health insurance coverage across the population through the Social Health Authority (SHA) to eliminate the financial access barriers to health and social development programs.^55^

Given the constitutionally sanctioned devolution in Kenya, national health outcomes now and in the future will inevitably be linked to county-level performance where elected leaders and technical bureaucrats make decisions on priorities, resource allocation and management.^56^ These regional units shoulder heavy responsibilities to achieving nationally set goals such as UHC with implications for national economic performance on which the county allocations depend. As such, the planning and implementation of health programs, including resource mobilization and re-allocation implications, must be measured by the need to balance the autonomy of devolved units and effectiveness of health delivery to meet population needs.^57^

Addressing scale and technical inefficiencies across the country is timely, particularly given the reduced fiscal space occasioned by the policies of the US government to cut donor assistance for health.^58^ ^59^ To facilitate shared learning and cross pollination of ideas across counties, it would be prudent to create opportunities and structures to share practical managerial information of what works and what does not. These could be thematically organized into communities of practice or evidence networks covering key aspects of the health system management, where county health systems could learn from one another. This could also offer an opportunity for benchmarking and tracking progress as a way of keeping accountability to meeting the county and national health objectives.^60^ ^61^

In interpreting the findings of this study, we recognize several limitations. We have had to rely on the most recent available data for the country to develop a consistent analytical framework. Some of these data were collected for different purposes and archived in various sources of variable quality. Further, we did not have data on private health spending as an input and only relied on the public health spending, assuming that this was the predominant source of health funding for most counties in Kenya. However, the inability to include the private health expenditure in our analysis could artificially inflate the efficiency of counties where private spending is significant, since we might have underestimated the input side of the equation. To mitigate for this potential limitation, we have included childhood immunization, which is mostly publicly funded, as a health output in our analysis, and the results are consistent with the overall analysis.

Another limitation is that we have considered health inputs and outputs for the same year of 2022, but we recognize that health investments tend to have timing lags to generate outputs. That is, an investment today affects outcomes tomorrow. However, it is reasonable to assume that health inputs to counties do not change significantly from those of the previous years. Lastly, we did not have data on prices of health inputs, which could have allowed us to estimate the allocative efficiency of county health systems.^21^ ^24–28^ However, we have made every effort to address those shortcomings by carefully curating the available data and applying robust analytical methods to arrive at consistent and informative results of beneficial policy impact. As more recent datasets become available, they can be regularly analyzed employing the same approach to deliver useful insights over time in order monitor progress and inform policy and implementation decisions at various levels.

## Conclusion

Based on experience from other countries that have made progress towards UHC and improved population health outcomes, it is well appreciated that health system decision-makers need to carefully plan and track resource utilization towards set goals and objectives. This often entails regular collection and collation of data to link health system inputs to population-level outcomes logically. This measurement approach should be comprehensive and focused on key policy areas and interventions to enhance its utility for decision-making at various health system levels.^62^

Our study is one such effort that critically examines health system performance in the backdrop of decentralization of health services by combining data from multiple sources into a consistent assessment framework. We conclude that ongoing decentralization efforts in Kenya must pay attention to the inappropriate scale of the health operations at the county level to perform optimally. These observations, we believe, lend critical insights to policy for the 2030 UHC agenda in Kenya and that lessons could be drawn for countries with similar circumstances.

## Data Availability

The main data sets supporting the conclusions of this article are available on request and with written permission from the Kenya National Bureau of Statistics. The data from IHME are publicly available and can be accessed through this publication:
https://www.thelancet.com/journals/langlo/article/PIIS2214-109X(18)30472-8/fulltext

## Authors’ contributions

TA conceptualized the study. NR, DB and JT collated the data and did the preliminary analysis. TA, YK, WO, MS and LW did the data analysis. TA and YK wrote the first draft and WO, LW, RW, TO, AL, UA, MS did the detailed review and provided comments.

## Acknowledgements

The authors are grateful to the Kenya National Bureau of Statistics that collected and shared some of the data that has been used in this project. Further, the authors appreciate the contribution of the Institute of Health Metrics and Evaluation, University of Washington, that provided the subnational data on health burden.

## Data Sharing

The main data sets supporting the conclusions of this article are available on request and with written permission from the Kenya National Bureau of Statistics. The data from IHME are publicly available and can be accessed through this publication: https://www.thelancet.com/journals/langlo/article/PIIS2214-109X(18)30472-8/fulltext

## Patient and Public Involvement

Patients and/or the public were not involved in the design, or conduct, or reporting, or dissemination plans of this research.

---

**Appendix 1:**
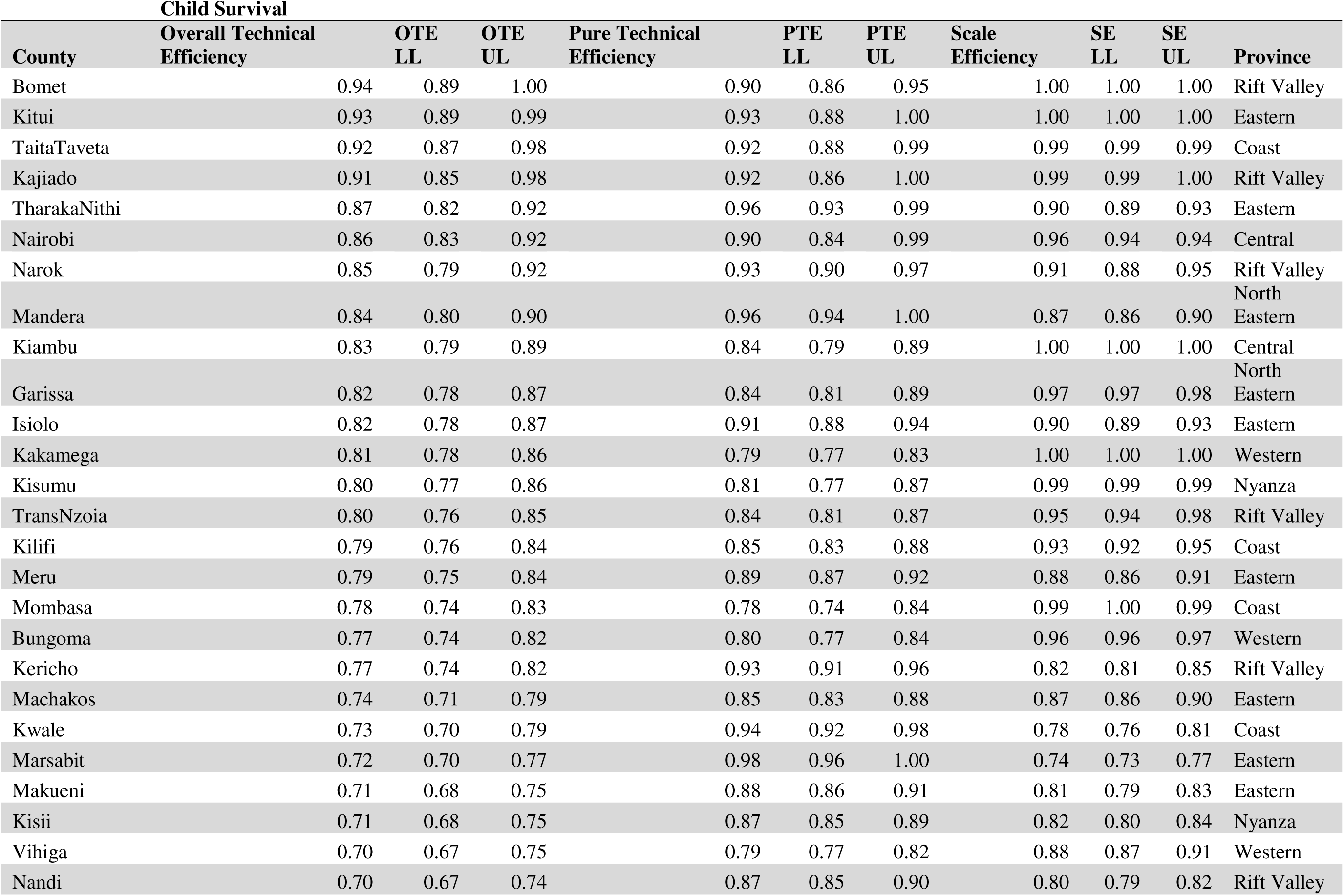
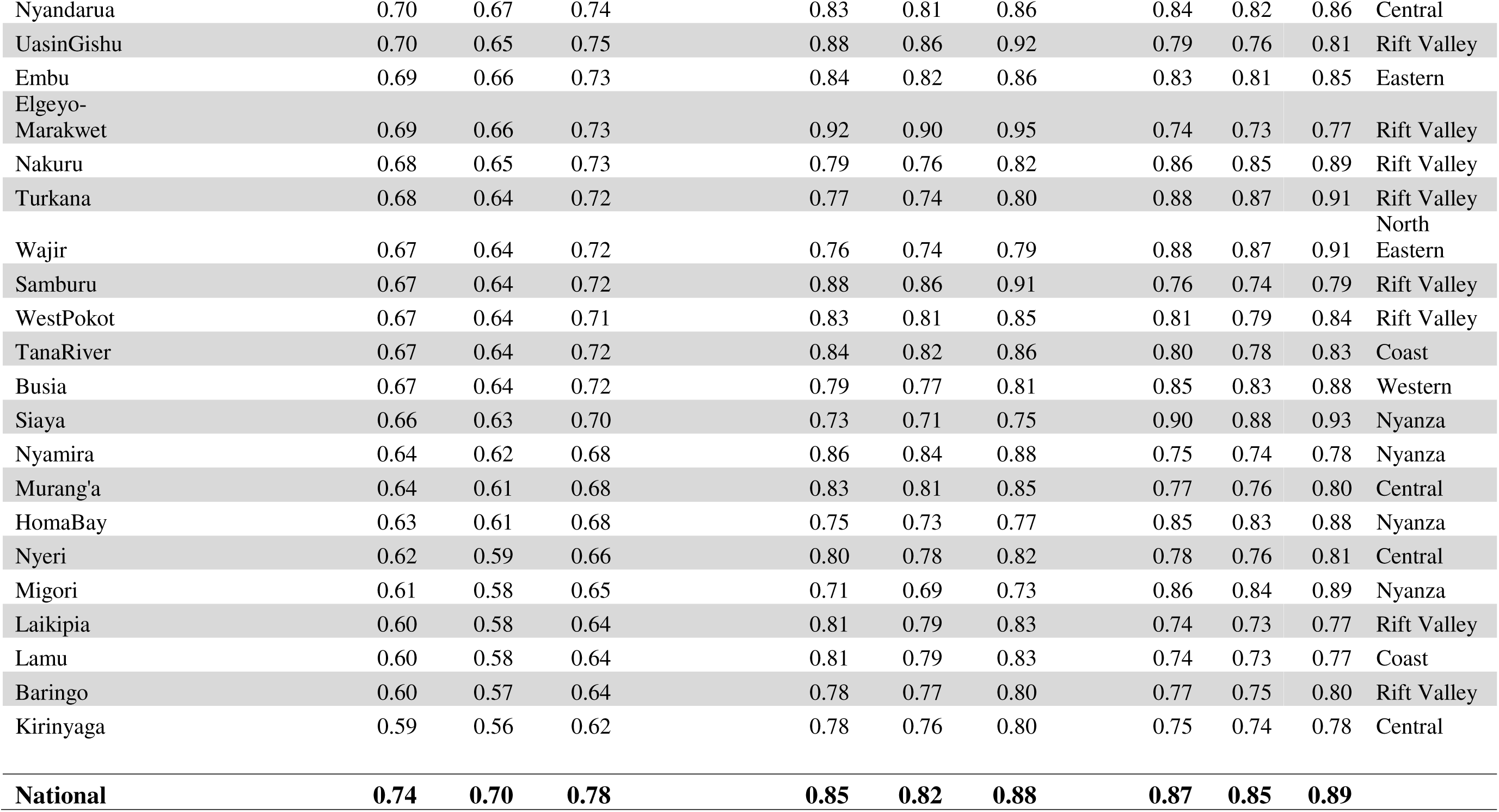
Child Survival.

**Appendix 2:**
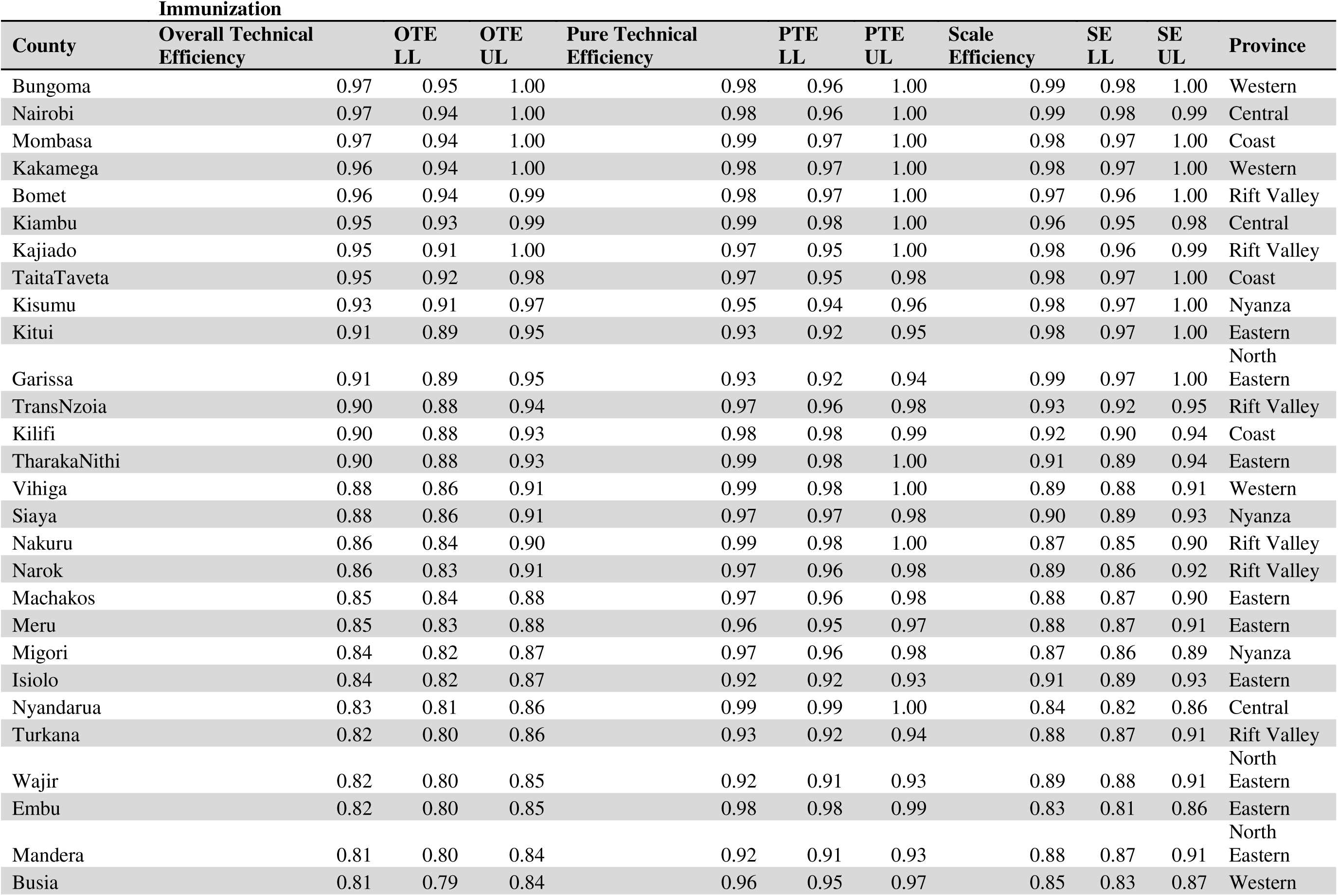
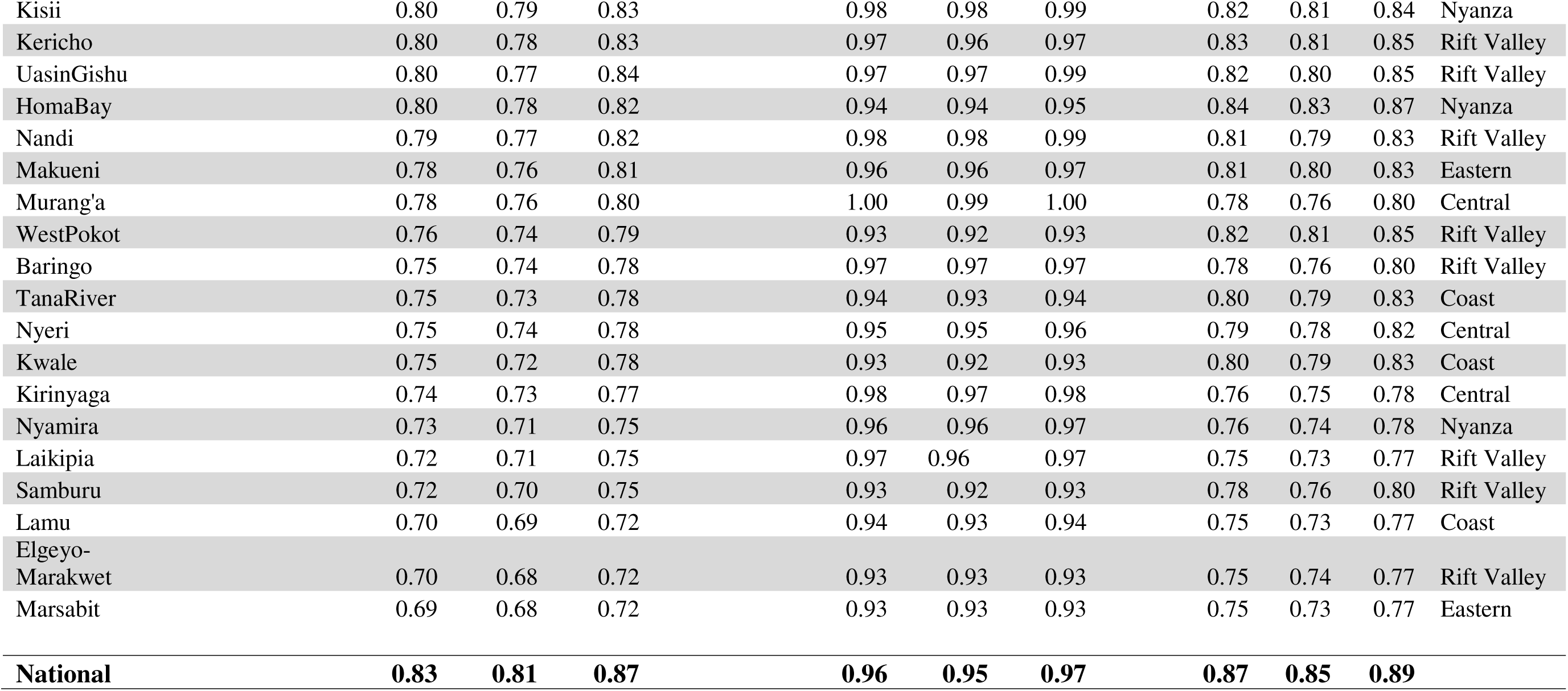
Immunization.

**Appendix 3:**
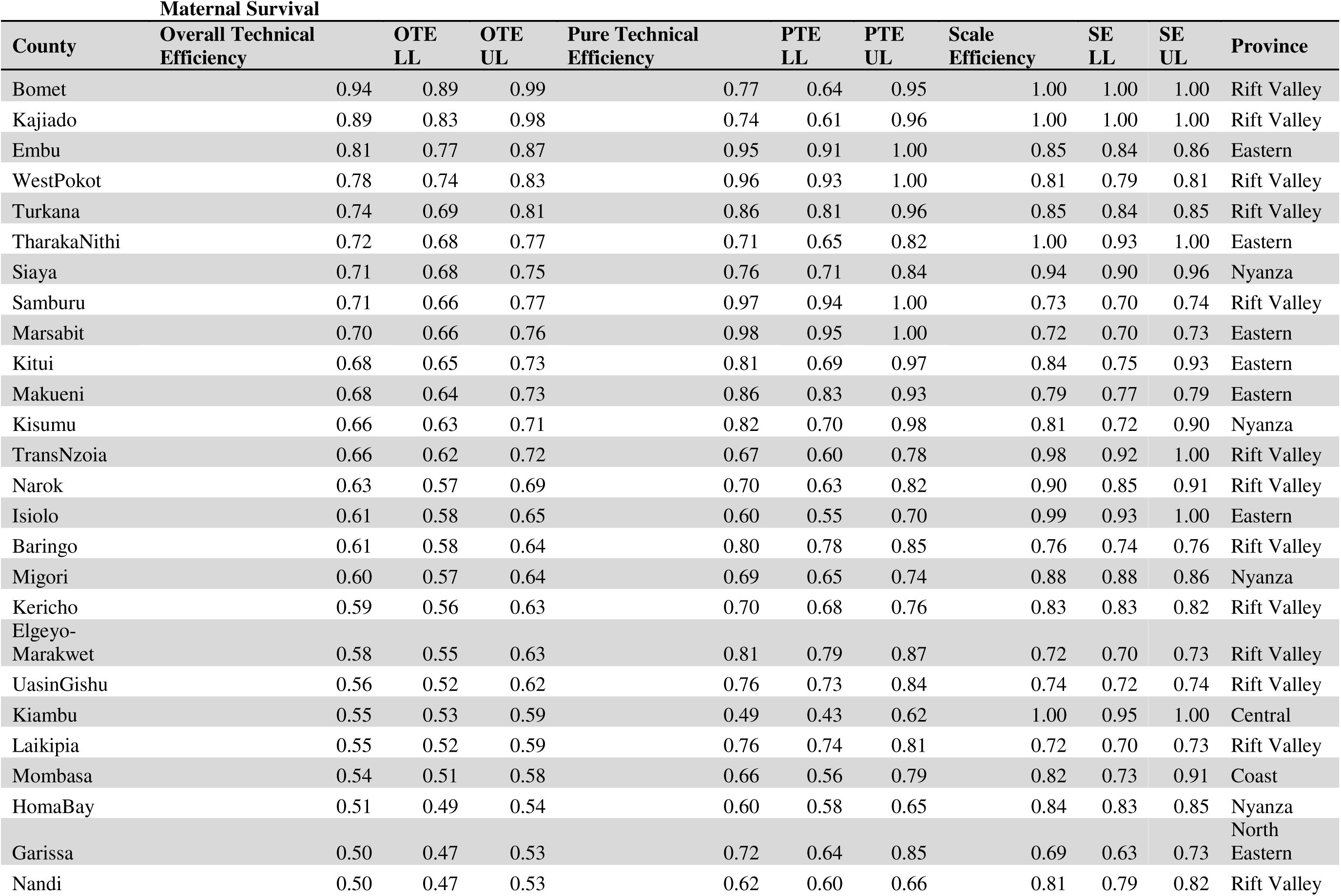
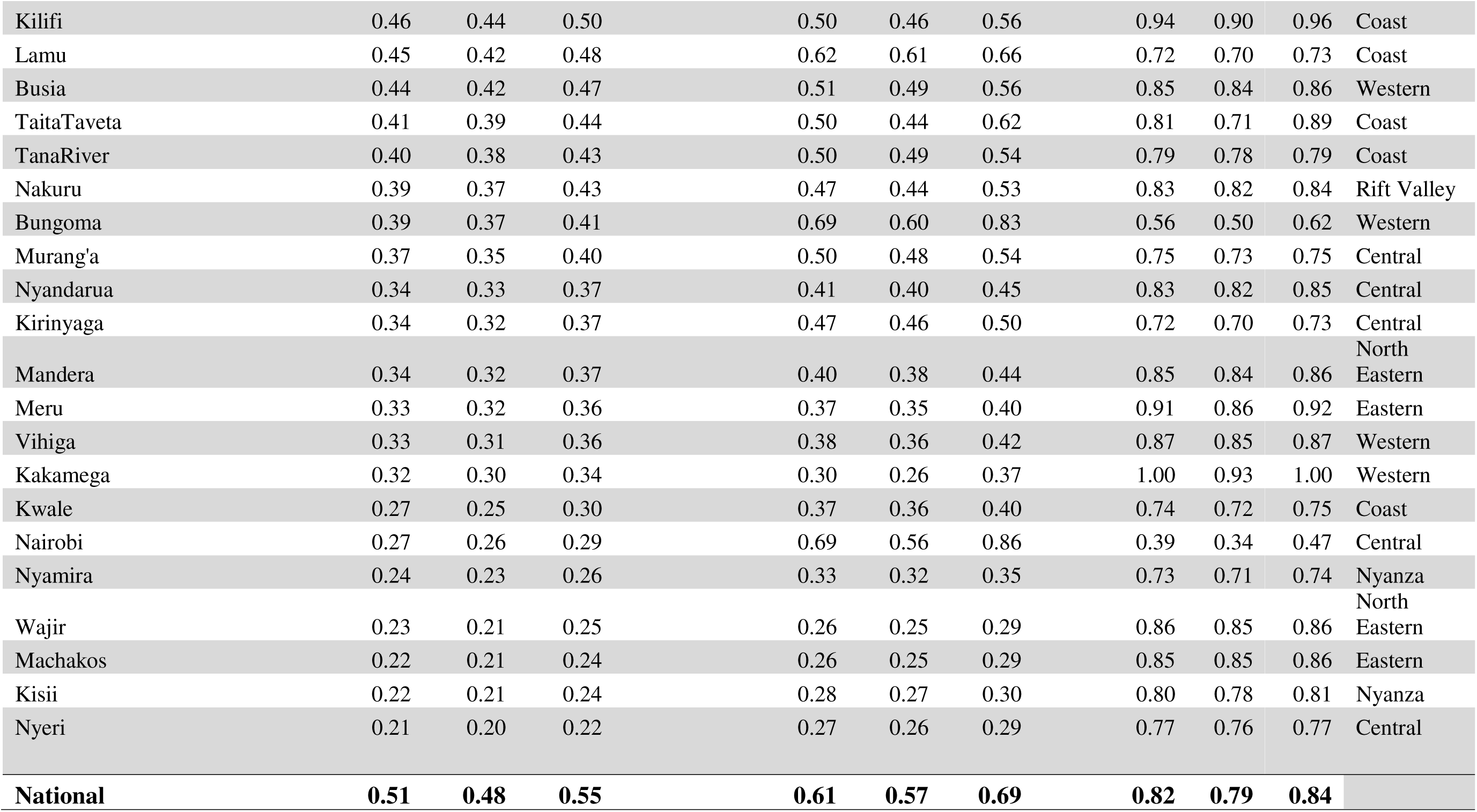
Maternal Survival.

**Appendix 4:**
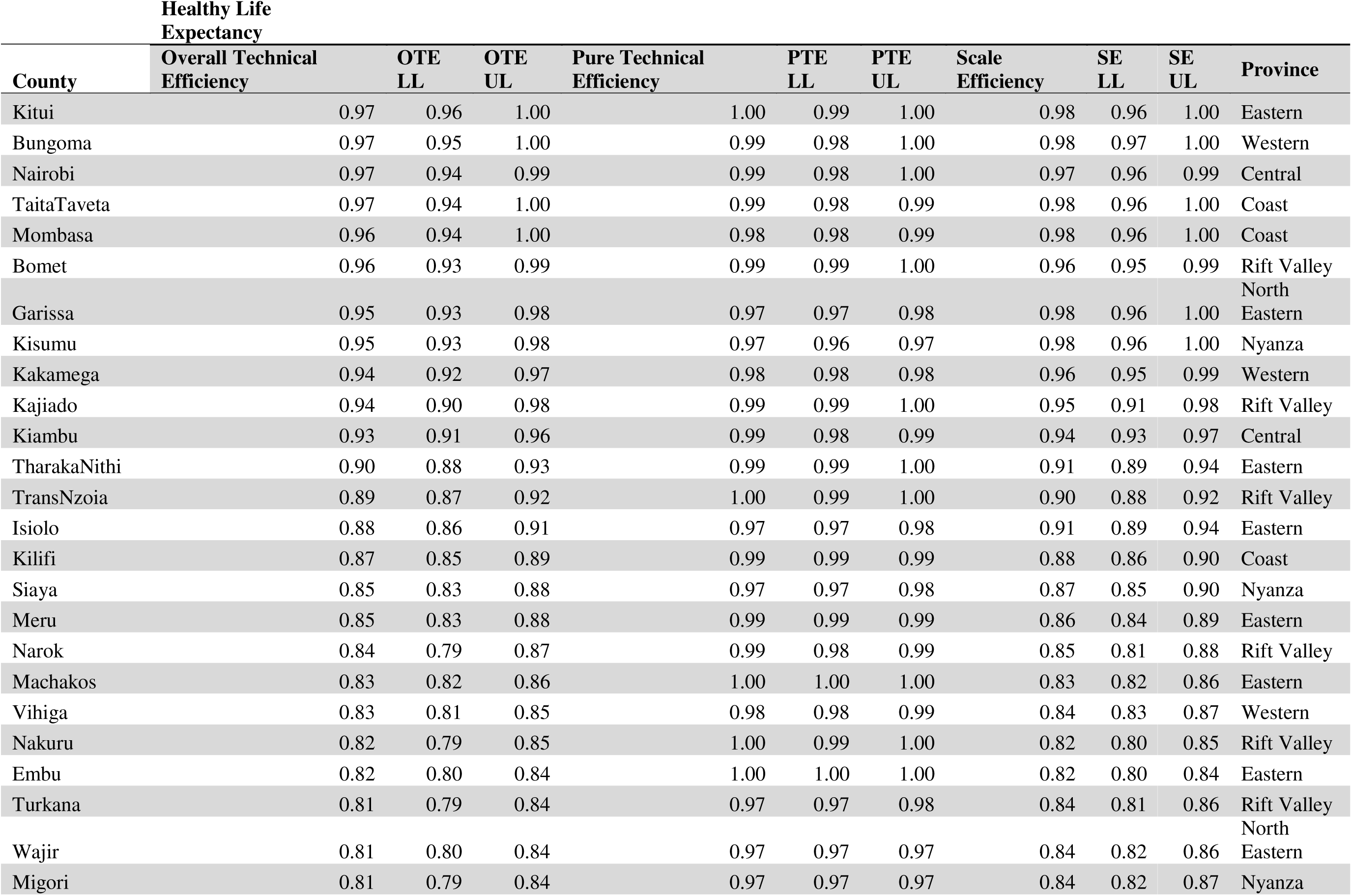
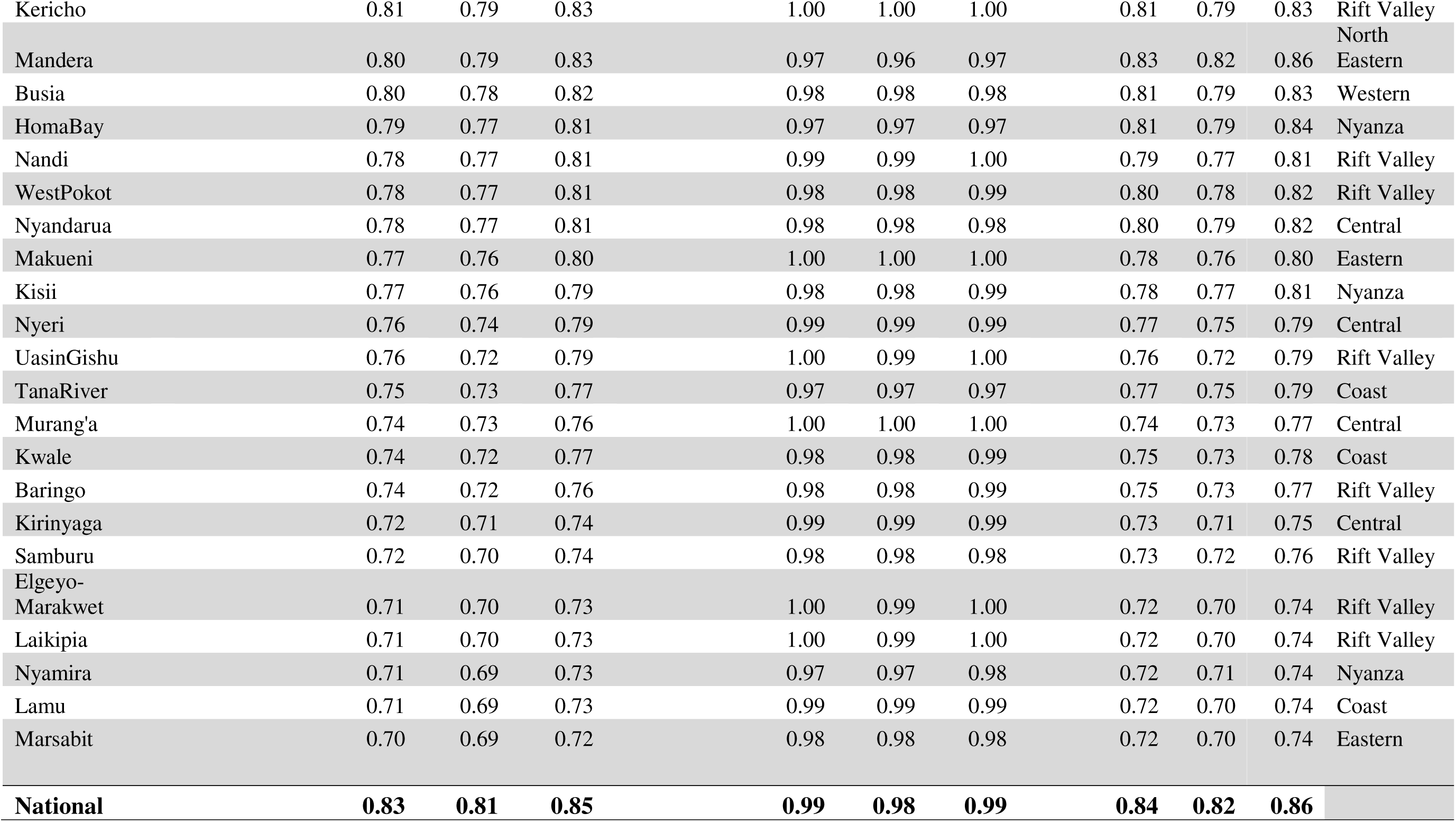
Healthy Life Expectancy.

**Appendix 5:** Provincial Ranking.

| **A.** | **Child survival** | **Overall Technical Efficiency** | **Standard Deviation** | **Minimum OTE** | **Maximum OTE** | **Counties** |
| --- | --- | --- | --- | --- | --- | --- |
|  | **Eastern** | 0.78 | 0.09 | 0.69 | 0.93 | 8 |
|  | **North Eastern** | 0.78 | 0.09 | 0.67 | 0.84 | 3 |
|  | **Coast** | 0.75 | 0.11 | 0.6 | 0.92 | 6 |
|  | **Western** | 0.74 | 0.06 | 0.67 | 0.81 | 4 |
|  | **Rift Valley** | 0.73 | 0.11 | 0.6 | 0.94 | 14 |
|  | **Central** | 0.71 | 0.11 | 0.59 | 0.86 | 6 |
|  | **Nyanza** | 0.68 | 0.07 | 0.61 | 0.8 | 6 |
| **B.** | **Immunization** |  |  |  |  |  |
|  | **Western** | 0.91 | 0.08 | 0.81 | 0.97 | 4 |
|  | **North Eastern** | 0.85 | 0.06 | 0.81 | 0.91 | 3 |
|  | **Central** | 0.84 | 0.10 | 0.74 | 0.97 | 6 |
|  | **Coast** | 0.84 | 0.12 | 0.7 | 0.97 | 6 |
|  | **Eastern** | 0.83 | 0.07 | 0.69 | 0.91 | 8 |
|  | **Nyanza** | 0.83 | 0.07 | 0.73 | 0.93 | 6 |
|  | **Rift Valley** | 0.81 | 0.08 | 0.7 | 0.96 | 14 |
| **C.** | **Maternal survival** |  |  |  |  |  |
|  | **Rift Valley** | 0.65 | 0.15 | 0.39 | 0.94 | 14 |
|  | **Eastern** | 0.59 | 0.21 | 0.22 | 0.81 | 8 |
|  | **Nyanza** | 0.49 | 0.21 | 0.22 | 0.71 | 6 |
|  | **Coast** | 0.42 | 0.09 | 0.27 | 0.54 | 6 |
|  | **Western** | 0.37 | 0.06 | 0.32 | 0.44 | 4 |
|  | **North Eastern** | 0.36 | 0.14 | 0.23 | 0.5 | 3 |
|  | **Central** | 0.35 | 0.12 | 0.21 | 0.55 | 6 |
| **D.** | **Hale** |  |  |  |  |  |
|  | **Western** | 0.89 | 0.08 | 0.8 | 0.97 | 4 |
|  | **North Eastern** | 0.85 | 0.08 | 0.8 | 0.95 | 3 |
|  | **Eastern** | 0.84 | 0.08 | 0.7 | 0.97 | 8 |
|  | **Coast** | 0.83 | 0.12 | 0.71 | 0.97 | 6 |
|  | **Central** | 0.82 | 0.11 | 0.72 | 0.97 | 6 |
|  | **Nyanza** | 0.81 | 0.08 | 0.71 | 0.95 | 6 |
|  | **Rift Valley** | 0.81 | 0.08 | 0.71 | 0.96 | 14 |

